# Role of Stress Echocardiography in the Assessment of Myocardial Ischemia

**DOI:** 10.1101/2023.09.18.23295624

**Authors:** Dhwani Manishbhai Patel, Binay K Panjiyar, Monica Ghotra, Gulam Husain Nabi Husain Mahato, Ronak Brijeshkumar Upadhyay, Abhimanyu Agarwal

## Abstract

Stress Echocardiography (ECG) is a commonly used modality for detection and assessment of Ischemic heart diseases (IHD). Its non-invasive nature makes it a more reliable diagnostic tool. This modality induces myocardial stress through exercise or pharmacological agents. Stress Echocardiography induced by exercise stress tests are more physiologic than pharmacologic stress tests as its finding tells about a patient’s exercise capacity which is prognostically important. Thus, if a patient can exercise, this is the preferred stress modality. Moreover, its radiation-free nature makes it a preferred option for individuals with contraindications to other stress imaging techniques and also decreases complications associated with other cardiac imaging modalities. Clinical conditions can be accurately assessed by comparing the findings of the heart rate and electrocardiograms after stress echocardiography with normal state. Analysis of stress echocardiography is done by visual precise evaluation of impaired myocardial contractility and regional wall motion abnormalities. This modality shows excellent results with current technology and by using an image enhancing agent, where necessary. It can identify the location of myocardial ischemia as well. Stress echocardiography holds significant potential in changing outcomes for a large population of patients with its high diagnostic accuracy, risk stratification capabilities, and cost-effectiveness.

## Introduction & Background

Myocardial ischemia, arising from an imbalance between myocardial oxygen supply and demand, is primarily caused by coronary artery disease (CAD), a prevalent condition characterized by atherosclerotic plaque formation in the coronary arteries. It is a significant cause of cardiovascular morbidity and mortality worldwide. Accurate assessment of myocardial ischemia is essential for guiding clinical decisions and implementing appropriate treatment strategies and timely management of condition. Stress echocardiography offers valuable insights into myocardial function and myocardial viability during stress.

Stress echocardiography involves combining echocardiographic imaging with physical (in the form of exercise) or pharmacological stress which allows clinicians to assess the heart’s response under challenging conditions. By inducing controlled stress, such as exercise or pharmacological agents, stress echocardiography can reveal abnormalities in myocardial blood flow and contractility, providing valuable information on the presence and severity of myocardial ischemia as well as the location of ischemia. It is also useful in preoperative evaluation for cardiovascular surgery and other major surgery and assessment of residual myocardial ischemia after revascularization procedures. Stress echocardiography provides real-time visualization of the heart’s response to stress, enabling the detection of regional wall motion abnormalities and alterations in left ventricular function, which are indicative of ischemia.

Cardiovascular disease results in 1 of every 3 deaths in the United States, or approximately 800 000 per year [1]. Among those who die suddenly of CHD, more than half have no antecedent symptoms [2]. In addition, myocardial infarction is frequently silent [3,4], causing no recognized symptoms but negatively affecting prognosis [3,4].

Several studies have demonstrated the diagnostic and prognostic utility of stress echocardiography in ischemic heart disease.While Stress echocardiography is an important method to diagnose coronary artery disease, it is based on the subjective assessment of changes in Left ventricle wall motion abnormality [5]. In the future, this obstacle may be overcome by the incorporation of artificial intelligence (AI) tools into the clinic capable of performing a quantitative assessment of stress images [6-8]. By examining coronary flow velocity reserve during stress echocardiography, researchers have identified functional and anatomical correlates that can aid in risk stratification and guide therapeutic decision-making [9].

To ensure the consistent and accurate interpretation of stress echocardiograms, guidelines have been established by professional societies like the American Society of Echocardiography. These guidelines offer valuable recommendations for the performance, interpretation, and application of stress echocardiography in clinical practice [10].

In real-world settings, stress echocardiography has demonstrated incremental diagnostic and prognostic value, leading to improved patient outcomes and risk assessment [11]. Moreover, practical guidance for the implementation of stress echocardiography in routine clinical practice has been provided, facilitating its seamless integration into patient evaluation and management [12].

This research aims to explore the role of stress echocardiography in the assessment of myocardial ischemia, its diagnostic accuracy, prognostic significance, and practical implementation in clinical practice. By analyzing and synthesizing the findings from various studies and guidelines, this research seeks to shed light on stress echocardiography’s versatile and informative role in the management of patients with suspected or known coronary artery disease.

## Review

### Methods

- This systematic review focuses on clinical studies concerning the role of Stress Echocardiography in assessment of myocardial Ischemia. We excluded animal studies and publications that only discussed the methodology of stress echocardiography without its correlation with myocardial ischemia. The review follows the guidelines for Preferred Reporting Items for Systematic Reviews and Meta-Analyses (PRISMA) [13] for 2020 in Figure *1* and only uses data collected from published papers, eliminating the need for ethical approval.

**Figure 1:**
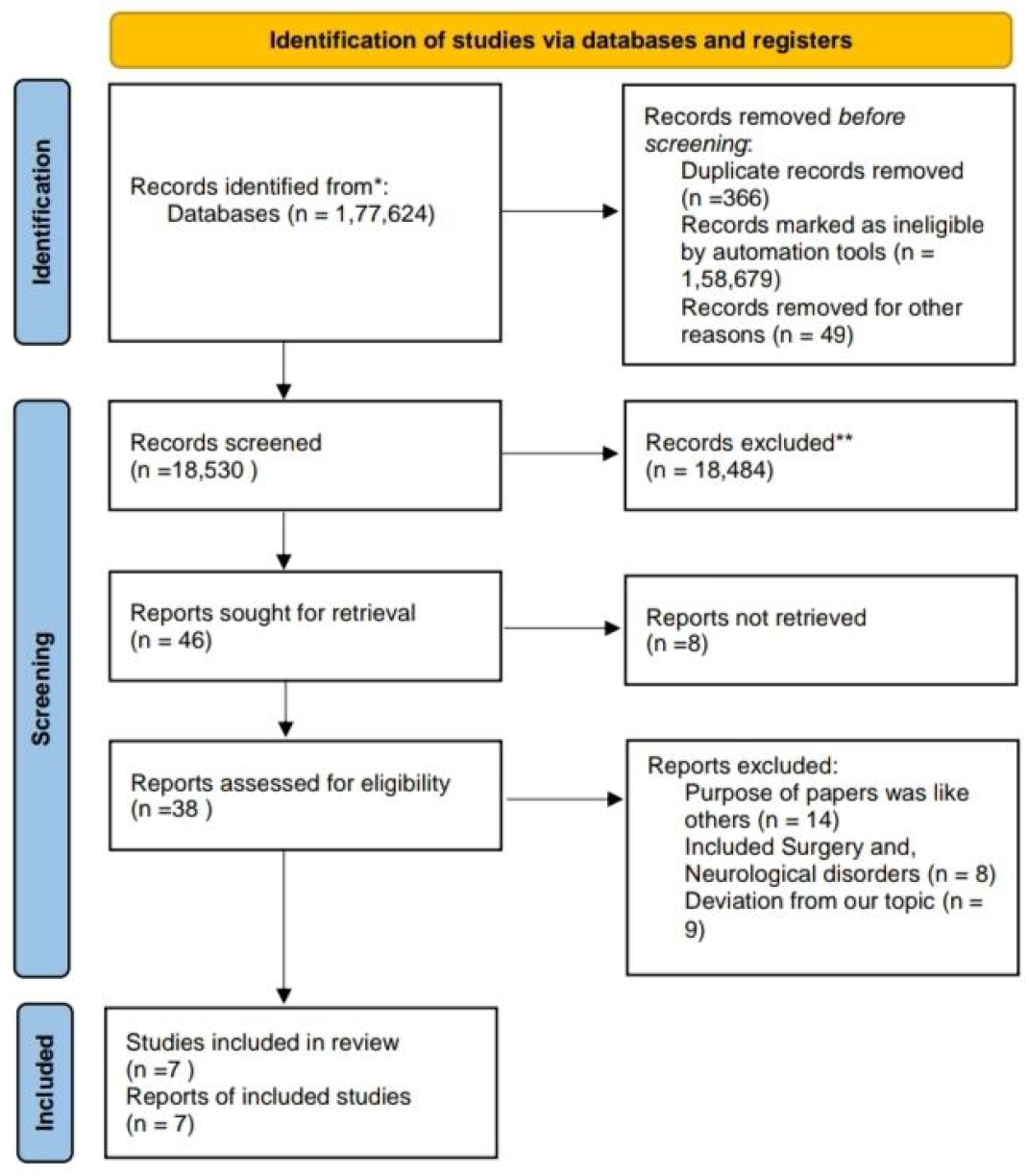
PRISMA flow diagram illustrating the search strategy and study selection process for the systematic review. PRISMA: Preferred Reporting Items for Systematic Reviews and Meta-Analyses

### Systematic Literature Search and Study Selection

We conducted a thorough search for relevant publications by using PubMed, including Medline and Google Scholar. We searched for studies mentioned in review papers, editorials, and commentaries on PubMed. Nevertheless, we continued searching for additional studies that satisfied our inclusion criteria.

We had a list of abstracts that we independently reviewed for inclusion using specific criteria. The criteria included association of stress echocardiography with myocardial ischemia or other ischemic heart diseases. We excluded review papers, animal studies and non English articles. Six reviewers conducted a dual review, and disagreements were resolved through discussion.

### Inclusion and Exclusion Criteria

We established specific criteria for including and excluding participants to achieve our study goals. Our Criteria can be summarized in Table 1.

**TABLE 1:** Showing the criteria adopted during the literature search process.

| Inclusion Criteria | Exclusion criteria |
| --- | --- |
| Studies on human | Studies on animal |
| Articles in English Language | Articles in Language other than English |
| From : 2013-2023 | Methodological studies explaining only stress echocardiography |
| Gender : All | Studies before 2013 |
| Age : >45 years | Age : <45 years |
| Free papers | Paid papers |
|  | Studies involving clinical data other than cardiovascular diseases |

### Search Strategy

The population, intervention/condition, control/comparison, and outcome (PICO) criteria were utilized to conduct a thorough literature review. The search was conducted on databases such as PUBMED (including Medline) and Google Scholar Libraries, using relevant keywords, such as stress echocardiography, myocardial ischemia and ischemic heart diseases. The medical subject heading (MeSH) approach for PubMed (including Medline) and Google Scholar, as detailed in Table 2, was employed to develop a comprehensive search strategy.

**TABLE 2:**
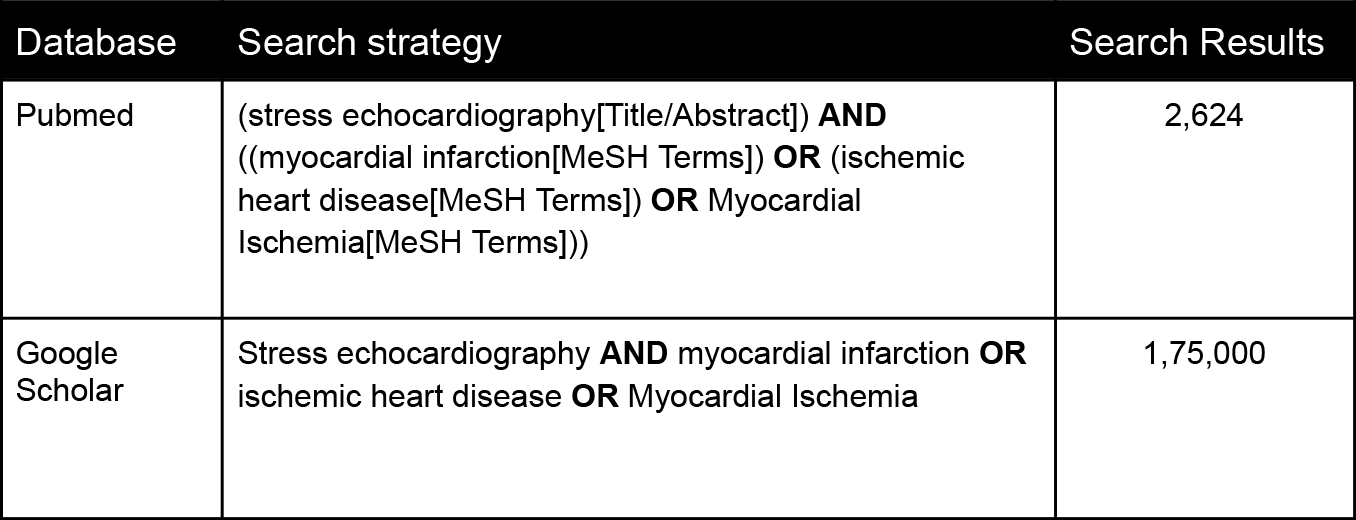
Showing the Search strategy, search Engines used, and the number of results displayed.

| Database | Search strategy | Search Results |
| --- | --- | --- |
| Pubmed | (stress echocardiography[Title/Abstract]) **AND** ((myocardial infarction[MeSH Terms]) **OR** (ischemic heart disease[MeSH Terms]) **OR** Myocardial Ischemia[MeSH Terms])) | 2,624 |
| Google Scholar | Stress echocardiography **AND** myocardial infarction **OR** ischemic heart disease **OR** Myocardial Ischemia | 1,75,000 |

### Quality Appraisal

To ensure the reliability of our chosen papers, we utilized various quality assessment tools. We employed the PRISMA checklist and Cochrane bias tool assessment for randomized clinical trials for systematic reviews and meta-analyses. Non-randomized clinical trials were evaluated using the Newcastle-Ottawa tool scale. We assessed the quality of qualitative studies, as shown in Table 3, using the critical appraisal skills program (CASP) checklist. To avoid any confusion in the classification, we utilized the scale for the assessment of narrative review articles (SANRA) to evaluate the article’s quality.

**Table 3:** Showing quality appraisal tools used. PRISMA: Preferred reporting items for systematic reviews and meta-analyses; SANRA: Scale for the assessment of non-systematic review articles

| Quality Appraisal Tools Used | Type of Studies |
| --- | --- |
| Cochrane Bias Tool Assessment | Randomized Control Trials |
| Newcastle-Ottawa Tool | Non-RCT and Observational Studies |
| PRISMA Checklist | Systematic Reviews |
| SANRA Checklist | Any Other Without Clear Method Section |

## Results

After searching through three selected databases, PubMed, Medline and Google Scholar, we extracted 177,624 articles. We then carefully reviewed each paper and applied specific criteria, which led to excluding 159,094 articles. From the remaining 18,530 papers, we chose not to utilize 18,484 of them due to duplicates or unsatisfactory titles and abstracts. We closely examined the remaining 46 papers and excluded 39 more as their content did not meet our inclusion criteria. Finally, we conducted a thorough quality check on the remaining 7 papers, which all met our criteria. These 7 articles are included in our final systematic review. Table 4 provides a detailed description of each.

**Table 4:**
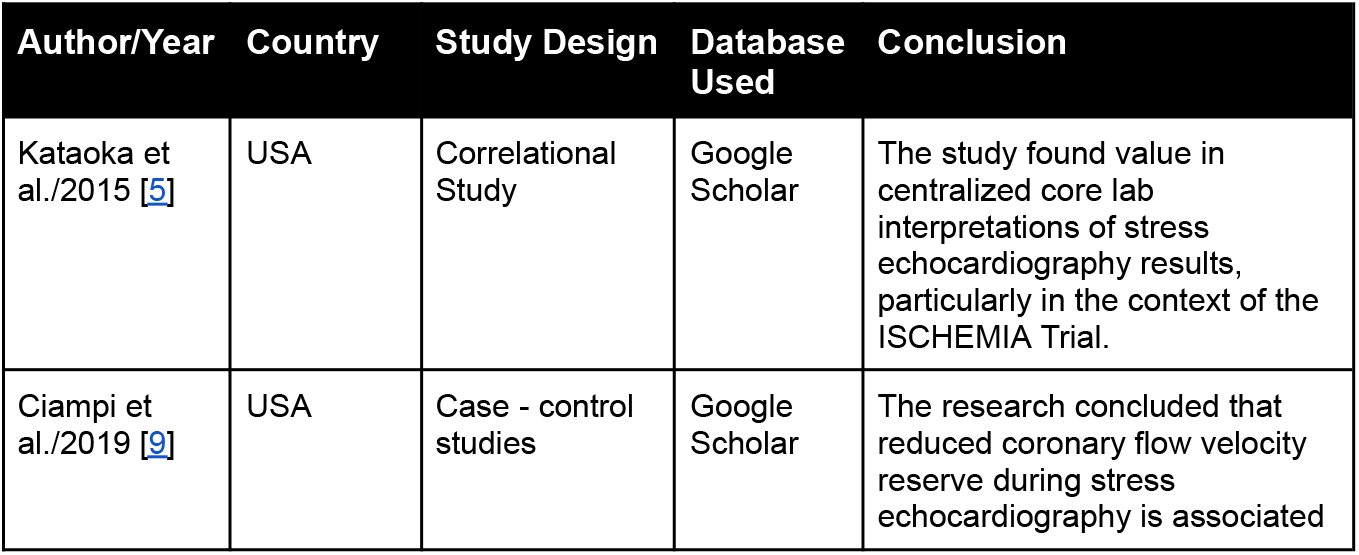
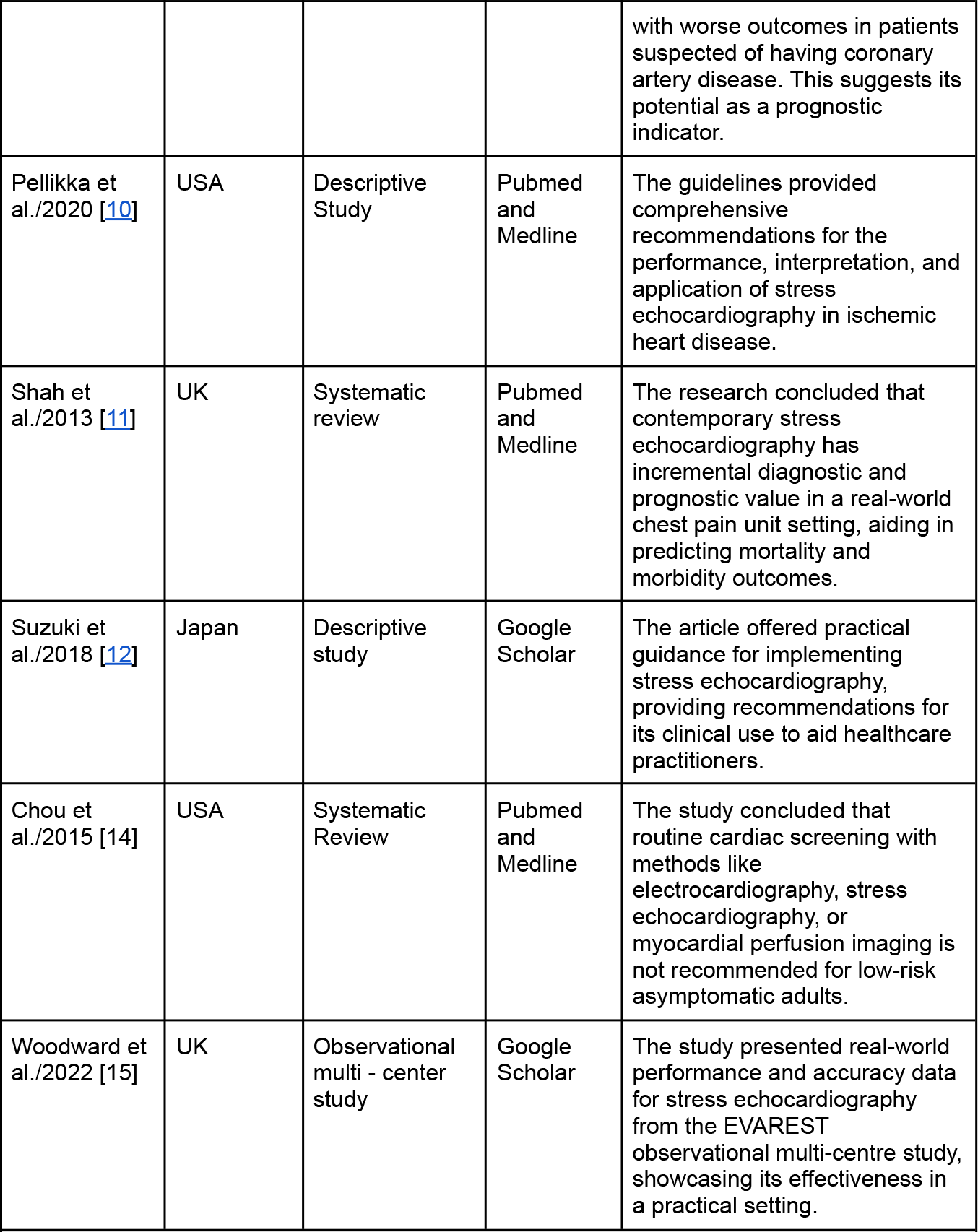
Summary of the results of the selected papers.

| Author/Year | Country | Study Design | Database Used | Conclusion |
| --- | --- | --- | --- | --- |
| Kataoka et al./2015 [5] | USA | Correlational Study | Google Scholar | The study found value in centralized core lab interpretations of stress echocardiography results, particularly in the context of the ISCHEMIA Trial. |
| Ciampi et al./2019 [9] | USA | Case - control studies | Google Scholar | The research concluded that reduced coronary flow velocity reserve during stress echocardiography is associated with worse outcomes in patients suspected of having coronary artery disease. This suggests its potential as a prognostic indicator. |
| Pellikka et al./2020 [10] | USA | Descriptive Study | Pubmed and Medline | The guidelines provided comprehensive recommendations for the performance, interpretation, and application of stress echocardiography in ischemic heart disease. |
| Shah et al./2013 [11] | UK | Systematic review | Pubmed and Medline | The research concluded that contemporary stress echocardiography has incremental diagnostic and prognostic value in a real-world chest pain unit setting, aiding in predicting mortality and morbidity outcomes. |
| Suzuki et al./2018 [12] | Japan | Descriptive study | Google Scholar | The article offered practical guidance for implementing stress echocardiography, providing recommendations for its clinical use to aid healthcare practitioners. |
| Chou et al./2015 [14] | USA | Systematic Review | Pubmed and Medline | The study concluded that routine cardiac screening with methods like electrocardiography, stress echocardiography, or myocardial perfusion imaging is not recommended for low-risk asymptomatic adults. |
| Woodward et al./2022 [15] | UK | Observational multi - center study | Google Scholar | The study presented real-world performance and accuracy data for stress echocardiography from the EVAREST observational multi-centre study, showcasing its effectiveness in a practical setting. |

## Discussions

Stress echocardiography (SE) is widely used for diagnosis, risk stratification, and prognosis of patients with known or suspected coronary artery disease and has reasonable sensitivity and specificity for clinical decision making [16,17]. “Screening” refers to testing for a disease or condition in asymptomatic persons to identify the condition before it manifests clinically [14]. It’s high diagnostic accuracy for ischemic heart diseases makes it preferable screening tool in real-world practice.

The Stress Echocardiography could be due to any mode of stress (exercise or pharmacologic) and could demonstrate any degree of ischemia from none to severe [5].During exercise to sustain the increased metabolic demand of the tissues, increased oxygen and nutrient delivery are accomplished by increasing cardiac output (CO) and blood flow to the microvascular surface area, in addition to increased O2 extraction in the myocardium. Examining cardiac function during exercise as well as at rest is thus important for selecting suitable therapy for heart disease [12]. Exercise stress testing has been widely undertaken for the diagnosis of heart diseases [18,19]. Stress echocardiography enlighten heart’s capacity to cope up with controlled stress.

In treadmill exercise, stress testing echocardiograms can be acquired after finishing the exercise. With supine ergometer exercise, stress testing echocardiograms can be acquired during exercise.In selecting the testing method, it is essential to give priority to the safety of the patient, taking into consideration his/her condition. For the elderly, cycle ergometer exercise is recommended as falls are unlikely and comparatively less workload in terms of oxygen consumption.Termination of exercise when the patient reaches the target heart rate or when any termination criterion is met (remarkable increase or decrease in blood pressure or significant arrhythmias); otherwise, continue up to the limit of tolerance of the patient. Patient condition and blood pressure must be monitored while using ECG and during exercise [12]. Tracking vitals of patients give approx idea of their capability of enduring stress.

In the circumstances where exercise stress cannot be performed, detailed evaluations under drug stimulation have been carried out to make an assessment of cardiac reserve. In such cases, dobutamine stress echocardiography(DSE) is indicated. This is a test to diagnose heart reserve by imposing stress on the heart with drug infusion. Dobutamine (a drug that stimulates the heart muscle) is used. This modality permits stable tomographic image recording during stress, allowing for more detailed assessment of Left ventricle (LV) wall motion, compared with exercise stress echocardiography. DSE is useful not only in the evaluation of myocardial ischemia, but also in the assessment of myocardial viability and contractile reserve in people following myocardial infarction or those with chronic Ischemic heart disease (IHD) [12]. In real-world setting, DSE is more preferable than Exercise induced stress echocardiography.

Being a test that imposes stress on the heart, stress echocardiography might result in the development of heart attack or arrhythmias, or very rarely, death [12]. Stress testing with vasodilators (dipyridamole or adenosine) may be performed for assessment of ischemia, myocardial perfusion, and myocardial viability [20.]. DSE rather than vasodilator stress echocardiography is preferred by most because of higher sensitivity for detection of Coronary artery disease (CAD) unless perfusion can also be assessed [10]. This modality is now being used in pediatric population too.

The baseline resting echocardiogram performed prior to initiation of stress should include a screening assessment of cardiac structure and function, including segmental and global ventricular function, chamber sizes, wall thickness, and cardiac valves, unless echocardiography has recently been performed. Standard views for assessment of regional wall motion and thickening include the parasternal long- and short-axis images and apical 4- and 2-chamber views [10]. The wall motion score index (WMSI) was calculated at rest and peak stress with the 16-segment model by adding the individual segment scores (1=normal, 2=hypokinesia, 3=akinesia, 4=dyskinesia) and dividing by 16.For assessment of regional myocardial function, either the 16- or 17 segment model of the LV may be used [21].In clinical practice in which regional wall motion (RWM) and thickening are assessed,the 16-segment model is commonly used. Rest and stress images were displayed side by side for ease of comparison [11]. Based on literature review and expert consensus, this was determined as occurring when at least 3 segments developed significant wall motion abnormality (WMA) during SE [22]. Thus, mild ischemia was defined as one or two segments with stress-induced WMAs [5].

Stress echocardiography (SE) is extensively utilized for diagnosing, assessing risk, and predicting outcomes in patients with confirmed or suspected coronary artery disease. It exhibits a reasonable degree of sensitivity and specificity for clinical decision-making [16,17]. The term “screening” pertains to testing asymptomatic individuals for a condition to identify it before clinical symptoms manifest [14]. Its high accuracy in diagnosing ischemic heart diseases makes it a preferred screening tool in real-world scenarios.

Stress echocardiography can be induced through exercise or pharmacological means, revealing varying levels of ischemia [5]. During exercise, the heart increases cardiac output and blood flow to meet heightened tissue demands, also extracting more oxygen in the myocardium. Evaluating heart function during both exercise and rest is crucial for determining appropriate therapies for heart conditions [12]. Exercise stress testing has been extensively used for heart disease diagnosis [18,19]. Stress echocardiography demonstrates the heart’s capacity to manage controlled stress.

In treadmill exercise, stress echocardiograms can be obtained post-exercise, while with supine ergometer exercise, they are acquired during exercise. Patient safety considerations guide the choice of testing method, with cycle ergometer exercise recommended for the elderly due to reduced fall risk and oxygen consumption workload. Exercise termination occurs at the target heart rate or when specific criteria are met, all while monitoring patient condition and blood pressure [12]. Monitoring vital signs provides an approximation of a patient’s stress tolerance.

When exercise stress isn’t feasible, drug-induced evaluations of cardiac reserve are conducted. In such cases, dobutamine stress echocardiography (DSE) is utilized. This test assesses heart reserve by stressing the heart with drug infusion, specifically dobutamine. DSE is valuable not just for identifying myocardial ischemia, but also for evaluating viability and contractile reserve in post-myocardial infarction patients or those with chronic ischemic heart disease [12]. In practical settings, DSE is often favored over exercise-induced stress echocardiography.

Stress echocardiography, while valuable, could potentially trigger heart attacks, arrhythmias, or rarely, death [12]. Vasodilator stress tests (using dipyridamole or adenosine) can assess ischemia, myocardial perfusion, and viability [20]. DSE is generally preferred due to its higher sensitivity in detecting Coronary Artery Disease (CAD), unless perfusion assessment is necessary [10]. DSE is now being extended to the pediatric population.

The baseline resting echocardiogram, conducted before stress, includes an assessment of cardiac structure and function. Evaluations involve ventricular function, chamber sizes, wall thickness, and cardiac valves. Standard views for assessing wall motion and thickening encompass various angles [10]. The Wall Motion Score Index (WMSI) is computed at rest and peak stress to gauge myocardial function [11]. Rest and stress images are compared side by side for analysis [11]. A significant wall motion abnormality (WMA) in at least 3 segments during SE indicates mild ischemia [22].

Stress echocardiograms are categorized as normal, abnormal ischemic, or abnormal nonischemic, each with distinct indications [11]. Monitoring wall motion during exercise helps determine the ischemic threshold. A resting regional wall motion abnormality reduces predictive accuracy for exercise and dobutamine stress echocardiography [15].

The number of LV wall segments with new wall motion abnormalities indicates the extent of ischemia, and the magnitude of abnormality reflects its severity. Both aspects should be evaluated under stress. Real-time myocardial contrast echocardiography (RTMCE) monitors blood flow changes during stress imaging [23]. AI integration could enhance reporting consistency and confidence, potentially expanding the range of personnel performing stress echocardiograms [15].

### Limitations

Our literature review has limitations as we limited our analysis to English articles on human studies published within the last 10 years, specifically targeting those who are more than 45 years old. We only used free articles and our study was limited to English papers on Stress echocardiography for myocardial ischemia and we excluded articles related to other modalities for assessment. We only assessed 7 articles related to our study. More research is needed for specific conclusions.

## Conclusion

Stress echocardiography is now being widely used for its reliability in assessing known or suspected Ischemic heart diseases. It is performed either by exercise (treadmill or bicycle) or in patients who are unable to do exercise; pharmacological agents (Dobutamine) or vasodilators (Adenosine) can also be used. Where necessary, Use of image enhancing agents give excellent results in assessment of abnormalities in the heart. This modality assesses myocardial ischemia, myocardial perfusion and myocardial viability too. It can detect LV dysfunction in the form of its wall motion abnormalities and can also tell about risk stratification and predict prognosis. It can also estimate the extent and severity of diseases. Interpretation of the results of stress echocardiography are done by trained physicians so the differences in this subjective assessment can be overcome by modern technology like AI. Overall, stress echocardiography is gaining popularity as a trusted & reliable assessment tool for myocardial ischemia and other ischemic heart diseases.

## Acknowledgements

PD made a major contribution to the article, such as the conception of the work and collection of data for the work, correction, tables, and figures editing, and drafted the manuscript from introduction to conclusion. BP contributes to collecting data, double checks for possible errors, and drafting the introduction and method section. MG participates in selecting data, checking for duplicated data, checking for possible errors, and participating in the drafting of method sections and tables. MGH participates in checking for data collection, references, and drafting the result section and discussion. RU participates in drafting discussions, data collection, checking for possible errors, and providing suggestions. AA contributes to abstract drafting, discussion editing, data collection, and checking for possible errors. MG participates in editing the abstract, providing. Suggestions, data collection, figure editing, and title modification. MGH and RU participates in data collection, checks for any possible errors, and drafts conclusions. AA participates in data collection and abstract editing, ensuring all guidelines are met, and drafts limitation sections. PD participates in generating ideas, providing suggestions, title modification, corrections, revising the manuscript, and drafting the introduction, method, and conclusion. All authors read and approved the final manuscript.

## References

1) Mensah GA, Brown DW. An overview of cardiovascular disease burden in the United States. Health Aff (Millwood). 2007;26(1):38–48. doi:10.1377/hlthaff.26.1.38

2) Go AS, Mozaffarian D, Roger VL, et al. Heart disease and stroke statistics--2013 update: a report from the American Heart Association [published correction appears in Circulation. 2013 Jan 1;127(1):doi:10.1161/CIR.0b013e31828124ad] [published correction appears in Circulation. 2013 Jun 11;127(23):e841]. Circulation. 2013;127(1):e6-e245. doi:10.1161/CIR.0b013e31828124ad

3) Sheifer SE, Gersh BJ, Yanez ND 3rd, Ades PA, Burke GL, Manolio TA. Prevalence, predisposing factors, and prognosis of clinically unrecognized myocardial infarction in the elderly. J Am Coll Cardiol. 2000;35(1):119–126. doi:10.1016/s0735-1097(99)00524-0

4) Sigurdsson E, Thorgeirsson G, Sigvaldason H, Sigfusson N. Unrecognized myocardial infarction: epidemiology, clinical characteristics, and the prognostic role of angina pectoris. The Reykjavik Study. Ann Intern Med. 1995;122(2):96–102. doi:10.7326/0003-4819-122-2-199501150-00003

5) Kataoka A, Scherrer-Crosbie M, Senior R, et al. The value of core lab stress echocardiography interpretations: observations from the ISCHEMIA Trial. Cardiovasc Ultrasound. 2015;13:47. Published 2015 Dec 18. doi:10.1186/s12947-015-0043-2

6) Alsharqi M, Woodward WJ, Mumith JA, Markham DC, Upton R, Leeson P. Artificial intelligence and echocardiography. Echo Res Pract. 2018;5(4):R115–R125. doi:10.1530/ERP-18-0056

7) Alsharqi M, Upton R, Mumith A, Leeson P. Artificial intelligence: a new clinical support tool for stress echocardiography. Expert Rev Med Devices. 2018;15(8):513–515. doi:10.1080/17434440.2018.1497482

8) Dey D, Slomka PJ, Leeson P, et al. Artificial Intelligence in Cardiovascular Imaging: JACC State-of-the-Art Review. J Am Coll Cardiol. 2019;73(11):1317–1335. doi:10.1016/j.jacc.2018.12.054

9) Ciampi Q, Zagatina A, Cortigiani L, et al. Functional, Anatomical, and Prognostic Correlates of Coronary Flow Velocity Reserve During Stress Echocardiography. J Am Coll Cardiol. 2019;74(18):2278–2291. doi:10.1016/j.jacc.2019.08.1046

10) Pellikka PA, Arruda-Olson A, Chaudhry FA, et al. Guidelines for Performance, Interpretation, and Application of Stress Echocardiography in Ischemic Heart Disease: From the American Society of Echocardiography. J Am Soc Echocardiogr. 2020;33(1):1-41.e8. doi:10.1016/j.echo.2019.07.001

11) Shah BN, Balaji G, Alhajiri A, Ramzy IS, Ahmadvazir S, Senior R. Incremental diagnostic and prognostic value of contemporary stress echocardiography in a chest pain unit: mortality and morbidity outcomes from a real-world setting. Circ Cardiovasc Imaging. 2013;6(2):202–209. doi:10.1161/CIRCIMAGING.112.980797

12) Suzuki K, Hirano Y, Yamada H, et al. Practical guidance for the implementation of stress echocardiography. J Echocardiogr. 2018;16(3):105–129. doi:10.1007/s12574-018-0382-8

13) Liberati A, Altman DG, Tetzlaff J, et al. The PRISMA statement for reporting systematic reviews and meta-analyses of studies that evaluate health care interventions: explanation and elaboration. J Clin Epidemiol. 2009;62(10):e1–e34. doi:10.1016/j.jclinepi.2009.06.006

14) Chou R; High Value Care Task Force of the American College of Physicians. Cardiac screening with electrocardiography, stress echocardiography, or myocardial perfusion imaging: advice for high-value care from the American College of Physicians. Ann Intern Med. 2015;162(6):438–447. doi:10.7326/M14-1225

15) Woodward W, Dockerill C, McCourt A, et al. Real-world performance and accuracy of stress echocardiography: the EVAREST observational multi-centre study. Eur Heart J Cardiovasc Imaging. 2022;23(5):689–698. doi:10.1093/ehjci/jeab092

16) Hennessy TG, Codd MB, Kane G, McCarthy C, McCann HA, Sugrue DD. Dobutamine stress echocardiography in the detection of coronary artery disease: importance of the pretest likelihood of disease. Am Heart J. 1997;134(4):685–692. doi:10.1016/s0002-8703(97)70052-8

17) Piérard LA, De Landsheere CM, Berthe C, Rigo P, Kulbertus HE. Identification of viable myocardium by echocardiography during dobutamine infusion in patients with myocardial infarction after thrombolytic therapy: comparison with positron emission tomography. J Am Coll Cardiol. 1990;15(5):1021–1031. doi:10.1016/0735-1097(90)90236-i

18) Pellikka PA, Nagueh SF, Elhendy AA, Kuehl CA, Sawada SG; American Society of Echocardiography. American Society of Echocardiography recommendations for performance, interpretation, and application of stress echocardiography. J Am Soc Echocardiogr. 2007;20(9):1021–1041. doi:10.1016/j.echo.2007.07.003

19) Lancellotti P, Pellikka PA, Budts W, et al. The clinical use of stress echocardiography in non-ischaemic heart disease: recommendations from the European Association of Cardiovascular Imaging and the American Society of Echocardiography [published correction appears in Eur Heart J Cardiovasc Imaging. 2017 May 1;18(8):832]. Eur Heart J Cardiovasc Imaging. 2016;17(11):1191-1229. doi:10.1093/ehjci/jew190

20) Picano E, Lattanzi F. Dipyridamole echocardiography. A new diagnostic window on coronary artery disease. Circulation. 1991 May;83(5 Suppl):III19-26. PMID: 2022042.

21) Lang RM, Bierig M, Devereux RB, et al. Recommendations for chamber quantification: a report from the American Society of Echocardiography’s Guidelines and Standards Committee and the Chamber Quantification Writing Group, developed in conjunction with the European Association of Echocardiography, a branch of the European Society of Cardiology. J Am Soc Echocardiogr. 2005;18(12):1440–1463. doi:10.1016/j.echo.2005.10.005

22) Shaw LJ, Berman DS, Picard MH, et al. Comparative definitions for moderate-severe ischemia in stress nuclear, echocardiography, and magnetic resonance imaging [published correction appears in JACC Cardiovasc Imaging. 2014 Jul;7(7):748]. JACC Cardiovasc Imaging. 2014;7(6):593-604. doi:10.1016/j.jcmg.2013.10.021

23) Wei K, Le E, Bin JP, Coggins M, Jayawera AR, Kaul S. Mechanism of reversible (99m)Tc-sestamibi perfusion defects during pharmacologically induced vasodilatation. Am J Physiol Heart Circ Physiol. 2001;280(4):H1896–H1904. doi:10.1152/ajpheart.2001.280.4.H1896

